# A Novel Approach to Data Driven Pandemic Recovery: The Pandemic Recovery Acceleration Model

**DOI:** 10.1101/2020.05.17.20104695

**Authors:** Jeffrey P. Gold, Christopher Wichman, Kenneth Bayles, Ali S. Khan, Christopher Kratochvil, James V. Lawler, John Martin Lowe, Shelly Schwedhelm, Brandon Grimm

**Affiliations:** Chancellor, University of Nebraska Medical Center and University of Nebraska Omaha; College of Public Health, University of Nebraska Medical Center; College of Medicine, University of Nebraska Medical Center; Global Center for Health Security, University of Nebraska Medical Center

## Abstract

A data driven approach to guide the global, regional and local pandemic recovery planning is key to the safety, efficacy and sustainability of all pandemic recovery efforts. The Pandemic Recovery Acceleration Model (PRAM) analytic tool was developed and implemented state wide in Nebraska to allow health officials, public officials, industry leaders and community leaders to capture a real time snapshot of how the COVID-19 pandemic is affecting their local community, a region or the state and use this novel lens to aid in making key mitigation and recovery decisions. This is done by using six commonly available metrics that are monitored daily across the state describing the pandemic impact: number of new cases, percent positive tests, deaths, occupied hospital beds, occupied intensive care beds and utilized ventilators, all directly related to confirmed COVID-19 patients.

Nebraska is separated into six Health Care Coalitions based on geography, public health and medical care systems. The PRAM aggregates the data for each of these geographic regions based on disease prevalence acceleration and health care resource utilization acceleration, producing real time analysis of the acceleration of change for each metric individually and also combined into a single weighted index, the PRAM Recovery Index. These indices are then shared daily with the state leadership, coalition leaders and public health directors and also tracked over time, aiding in real time regional and statewide decisions of resource allocation and the extent of use of comprehensive non-pharmacologic interventions.

## Background

During the current COVID-19 pandemic it is important that local and state health departments and coalitions are using actionable and accessible data to guide decision making and focus strategies. Also important is the collaboration of multiple sectors working together to develop real time data metrics^1,2^. This paper describes a data system that is assisting Nebraska’s six Health Care Coalitions (HCC) and 19 local health departments (LHD) with making decisions about relaxing or continuing their directed health measures. In addition, the paper illustrates the collaborative data sharing between health care, the states’ health information exchange, the state and local public health departments, and academe.

In late February of 2020, the University of Nebraska Medical Center (UNMC) Global Center for Health Security^3^ (GCHS) in collaboration with infectious disease experts, emergency management professionals and others worked to define a set of six commonly available metrics that would represent a daily snapshot of the COVID-19 progression / severity and the utilization of critical community health care resources. The relationships among this set of metrics developed, called the Pandemic Recovery Acceleration Model (PRAM), are computed on a daily basis from three disease and three resource specific values and are scalable from the level of an individual hospital up to a county, region or statewide status. The three disease specific values^4^ are the daily totals of new cases, newly reported deaths and the percent of new positive tests per day. The three health care resource specific values^5^ are the daily total of occupied acute care hospital beds, intensive care beds and ventilators in-use by confirmed COVID-19 positive patients per day. Each of the six values is associated with a regional and statewide per capita benchmark^6^.

## Metrics and Computations

The rolling average for two points in time, the immediate past 1 -3 days (M1) and the past 8-14 days (M2), are computed from each of the six values described above. The ratio (M3) of the two rolling means, current 3 days over the past week, is then calculated for each of the six reported values. Each of these metrics has a spatial analog: M1 is analogous to current position; M2 is analogous to past position and M3 is analogous to velocity. The three day interval was selected to smooth the impact of weekend access and reporting as well as to minimize large swings without artificially underestimating the current value; the prior 8-14 day, seven day interval, was selected to reflect a full cycle of pandemic spread from time of infection through screening, testing and reporting.

The computation of metrics M1, M2 and M3 are formalized in equations 1, 2 and 3. The capital letter M represents the metric computed based on either C (new cases), D (new deaths), T (percent positive tests), H (hospital beds occupied by patients with the disease), I (intensive care beds occupied by patients with the disease) or V (ventilators in use by patients with the disease). The lower case m represents a daily observed value and t represents the day for which the metric is being computed. Our work originally specified a three day average for M_1_; however, after peer review, it was suggested that M1 be variable and based on the average of a five day or even a full week average. Thus equation 1, formally acknowledges this suggestion. For the purposes of this report, t in equation 1 is 3 and greater and i = 0, 1, 2.

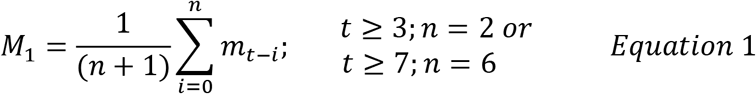

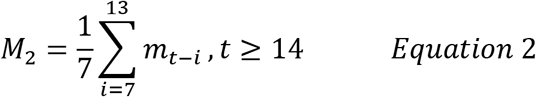

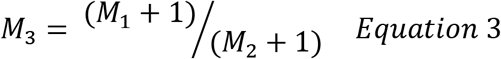

The addition of 1 to the denominator in equation 3 prevents division by 0; the addition of 1 in the numerator is to offset the 1 in the denominator.

Each of the velocity metrics (M3) are combined into four PRAM descriptive indices referred to as the Disease Trend Index (DTI), Resource Trend Index (RTI), Recovery Composite Index (RCI) and Recovery Ratio Index (RRI). Each index is analogous to the velocity and acceleration of change over the 14 day interval in a three dimensional axis defined by the changes of each of the three values (Figure 1). The DTI and RTI (Equations 4 and 5, respectively) are weighted averages of the disease specific velocities (C3, D3 and T3) and the resource specific velocities (H3, I3, V3), respectively. For the purposes of this paper, the weights for DTI and RTI were set to 1.

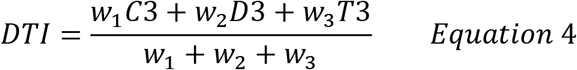

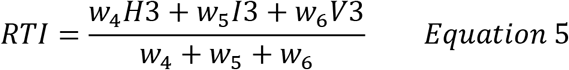

**Figure 1:**
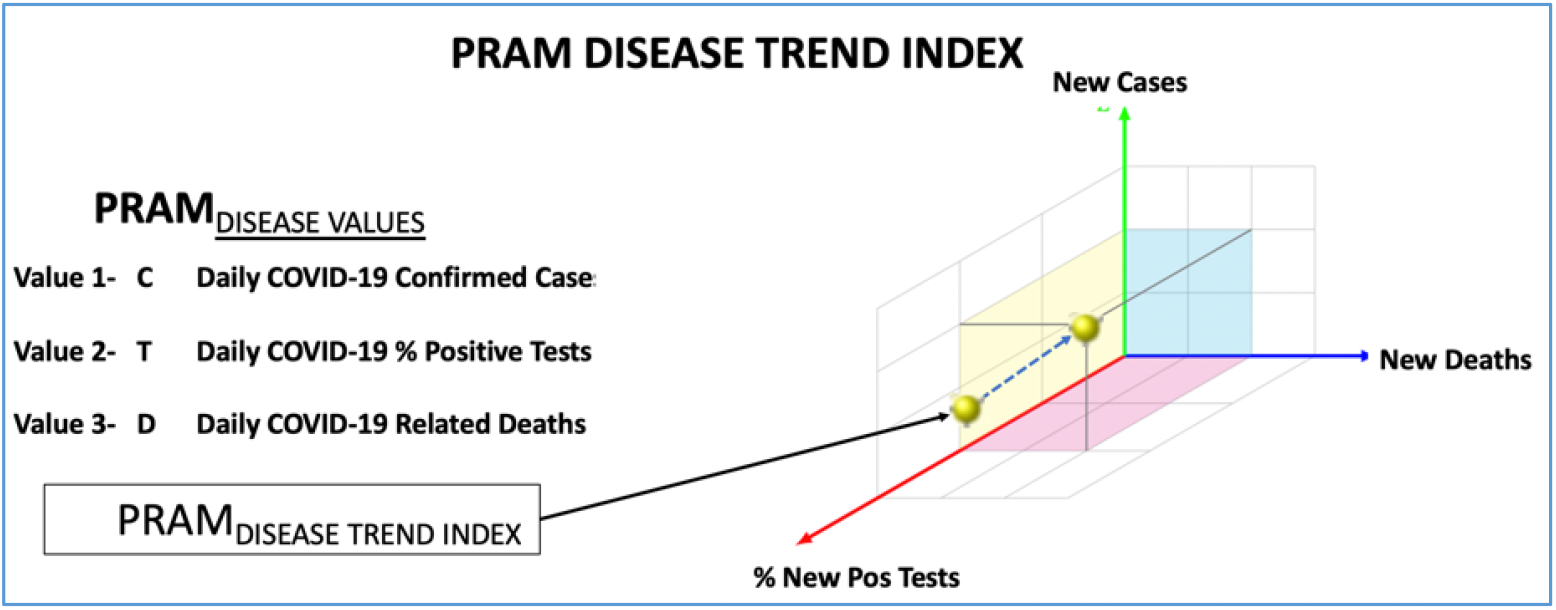
Three Dimensional Representation of Disease Trend Index

The DTI gives users a measure of regional or statewide disease spread and severity. For example, a DTI less than 1 implies all disease specific velocities are less than 1 and thus current new cases, new deaths and percent positive tests are all lower than the previous week. A DTI less than 1 for a sustained period of time would indicate the disease is receding (decelerating); a DTI equal to 1 implies all of the disease specific velocities are holding steady (not accelerating or decelerating); and a DTI greater than 1 indicates at least one of the disease specific velocities is greater than the previous week and the disease is progressing (accelerating). RTI is interpreted in a similar fashion as DTI, but for resource specific metrics.

The RCI is the average of the acceleration of the disease and the acceleration of the resource trend (RCI = ½ DTI + ½ RTI); providing a single value representing the acceleration or deceleration of pandemic recovery for the region. RCI is a less granular view of the pandemic with a similar interpretation to DTI and RTI. The RRI is the ratio of the acceleration of impact on health care resources divided by the acceleration of the impact of the pandemic spread velocity (RRI = RTI / DTI). Being a ratio of two severity indices, an RRI that is too small or too large is bad. For example, an RRI that is close to 0 would imply, resources may be available, but that disease spread is large. Since hospitalization tends to lag new cases, a small RRI may indicate that health care resources may soon be in demand. An RRI that is too large would imply that healthcare resources are being stretched even though the disease severity may be waning. These two latter indices will be particularly important as nonpharmacological interventions are relaxed and even more so when effective vaccines become available.

Plotting each metric and each index vs time yields a visual history of the disease impact and simultaneous resource utilization; the magnitude of each quantity is binned into three levels color coded as red (≥ 2), yellow (1 ≤ Index < 2) or green (< 1). Red indicates a rapid rate of rise (acceleration) in disease progression (DTI), resource utilization (RTI) or both combined (RCI), yellow indicates a slow rate of rise in disease progression or resource utilization and green indicates a slow rate of decrease (deceleration) in disease progression or resource utilization.

In addition to the running trends that provide the visual history of the metrics and indices, the GCHS also provides a daily color coded statewide and regional dashboard. A moment in time snapshot of the status of the state (Figure 2) and each of the geographic regions. The daily dashboard has seven columns; the first identifies the metric or index. The PRAM Today Benchmark column specifies the phase one benchmark value related to pandemic elimination. The benchmark values for the first six rows are set as follows:

**Figure 2:**
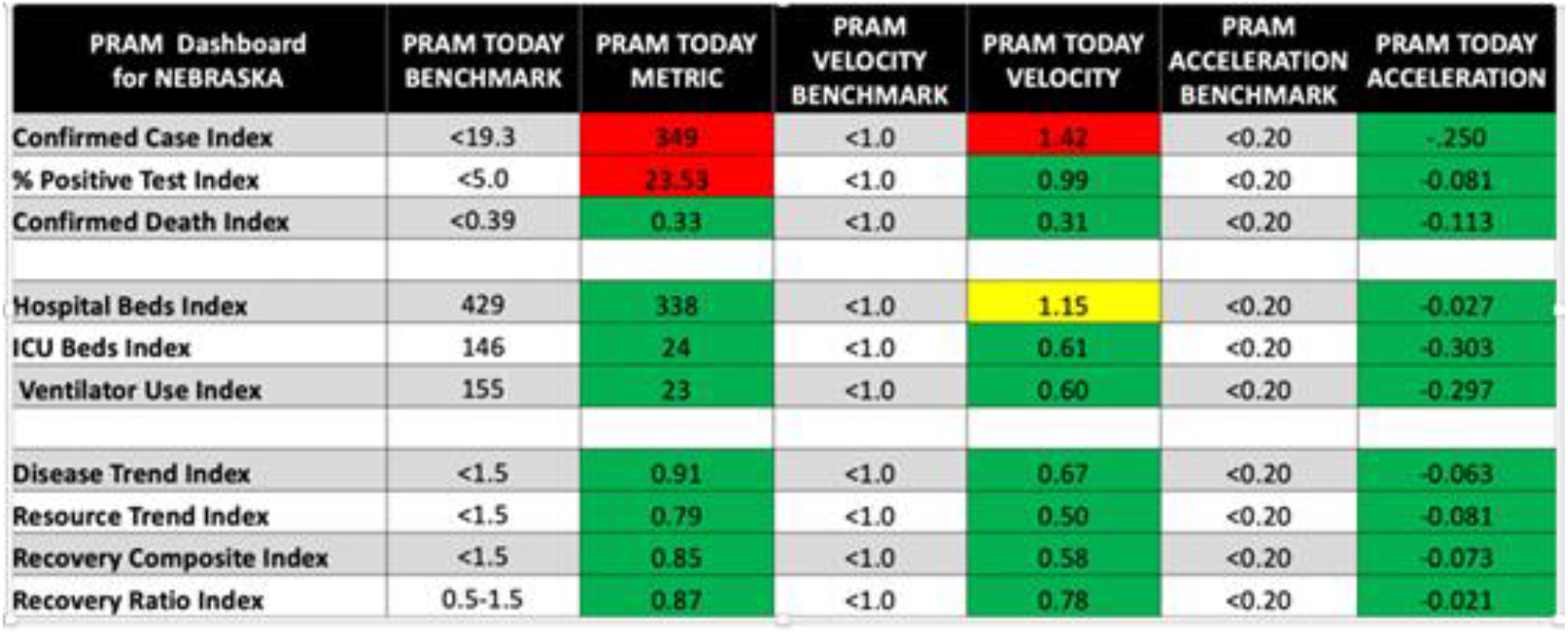
Global Center for Health Security PRAM Dashboard

1. CCI: 10 or fewer new cases per day per million population(pre-elimination)^6^;
2. PPTI: 5.0% positive diagnostic tests per day^i^;
3. CDI: 0.25 or fewer new deaths per day per million population^ii^;
4. HBI: 15% regional hospital bed capacity committed to COVID-19 patients^7^;
5. IBI: 20% regional intensive care bed capacity committed to COVID-19 patients^8^;
6. VUI: 25% regional ventilator care bed capacity committed to COVID-19 patients^iii^.

The benchmarks given for DTI, RTI, RCI and RRI were empirically derived over approximately six weeks of following the PRAM. The entries to the PRAM Today column are the current days’ M1 (new cases to ventilator care beds), DTI, RTI, RCI and RRI. The PRAM Velocity Benchmarks are the same as the colored bins used for the individual metric plots. The first six entries to the PRAM Today Velocity column are the current days’ M3 (new cases to ventilator care beds). The velocity for DTI, RTI, RCI and RRI is computed by applying Equation 3 to the respective daily index values. The PRAM Acceleration benchmarks were again set empirically after following the PRAM over a period of six weeks. The PRAM Today Acceleration is computed by dividing the difference between the current velocity and the velocity ten days prior by 10 (Equation 6).

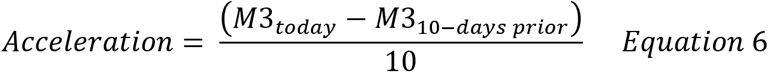

In the numerator of the acceleration, M3_10-days prior_ was chosen as it represents the velocity during the midpoint of the week in the denominator of M3_today_. The denominator of Equation 6 is the temporal difference between the current and 10-day prior velocities.

## Application of the PRAM to Nebraska

Nebraska’s six HCC’s are geographically distinct based upon the distribution of key health care resources, clinical care referral patterns, and they incorporate 19 local health departments (LHD) ^9^ Data for the disease metrics were obtained daily from the Nebraska Department of Health and Human Services Nebraska Electronic Disease Surveillance System (NEDSS); the resource metrics were obtained via the Nebraska Knowledge Center (KC) incident management software. KC is a situational intelligence platform for maintaining and managing operational control and creating a shared interoperable status in times of crisis. NEDSS data are aggregated counts by county per day; KC data are aggregated counts by HCC and LHD per day. The six metrics are aggregated on a daily basis for each of the six HCC’s and the state and weekly for each of the LHDs. The metrics are used to create a statewide (Figure 3) and a regional (Figure 4) PRAM dashboard. For the previous month, the figures above illustrate that the fourteen-day DTI acceleration for the state has varied from 1.2 to 2.5, while the acceleration of the DTI for the regional HCC illustrated has varied from 0.6 to a recent high of 9.5. This latter acceleration in this rural Nebraska HCC has been attributed to pandemic spread in high density organizations and residential communities. However, due to the low population density of this rural HCC, while it clearly has required immediate attention, it has impacted the statewide acceleration index only minimally over the last month PRAM analysis. This illustrates the importance of regional PRAM tracking in states with significant geographically dispersed rural and urban communities.

**Figure 3:**
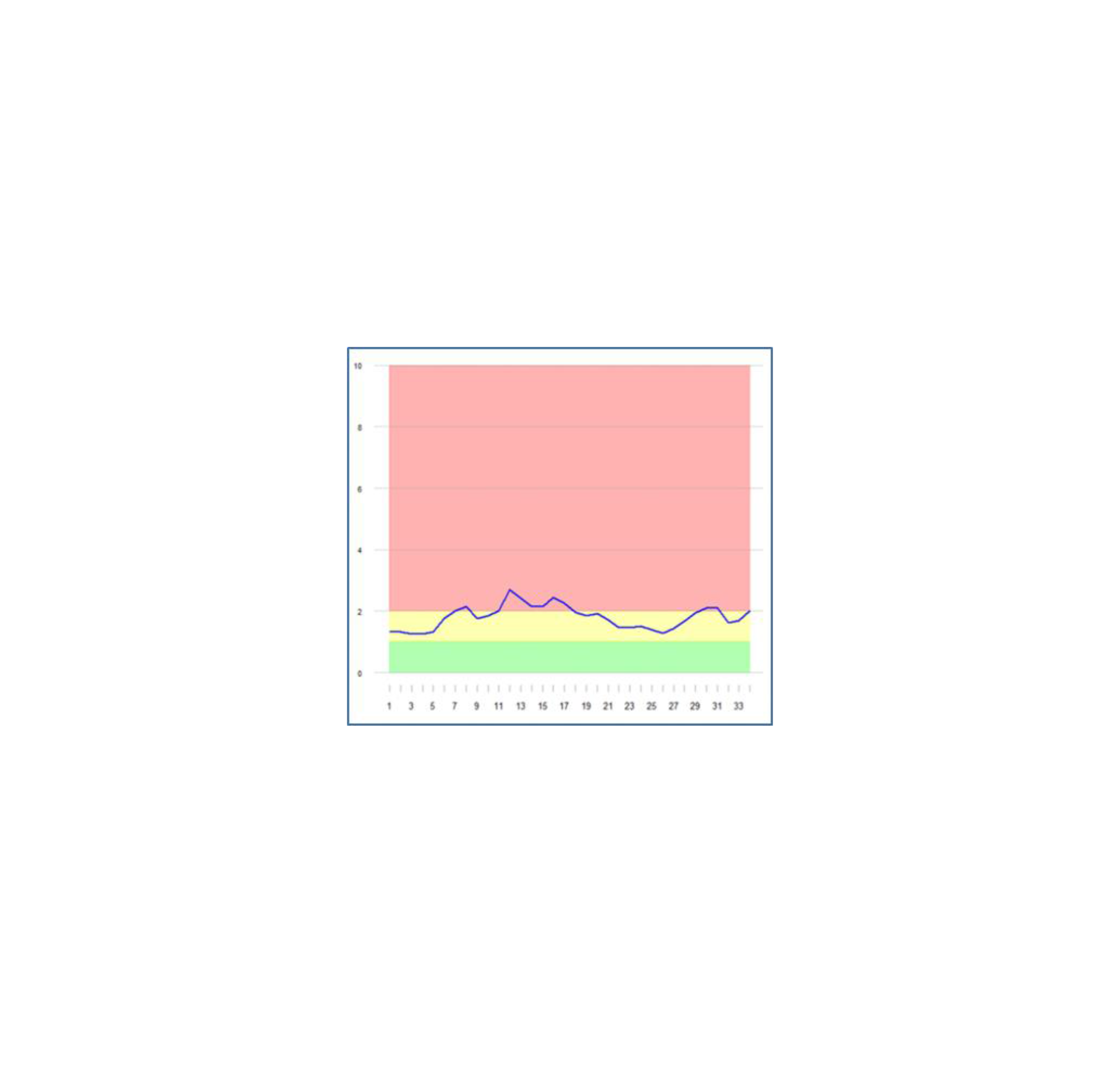
Statewide PRAM Disease Trend Index

**Figure 4:**
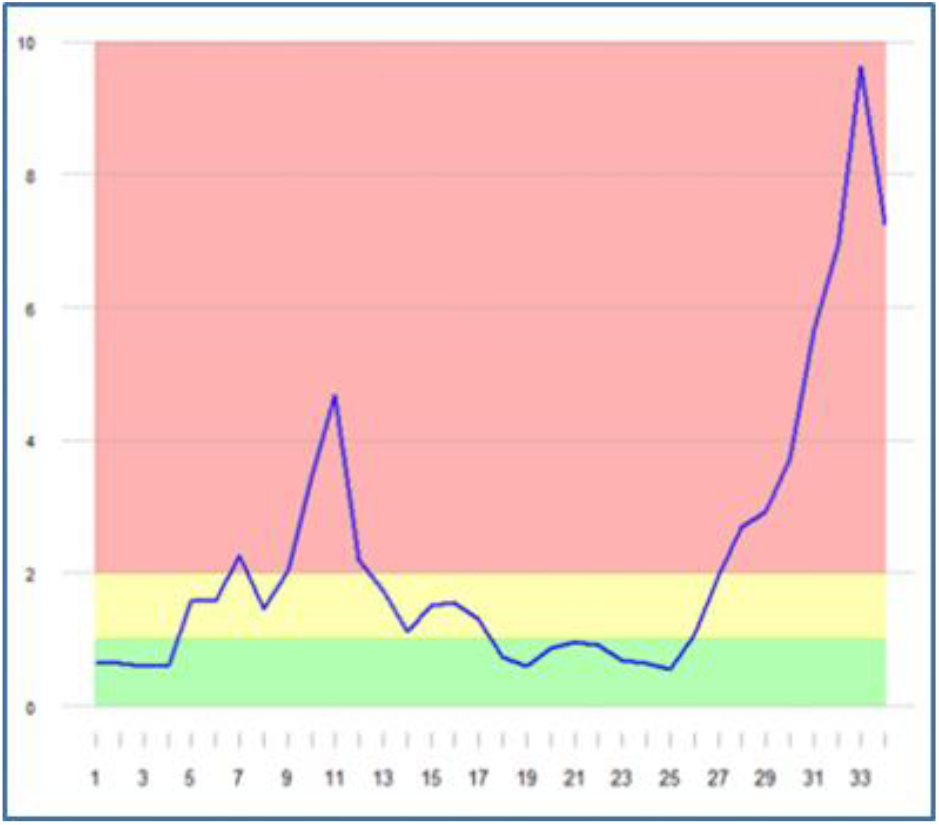
Regional PRAM Disease Trend Index

## Discussion

These aggregated, weighted acceleration indices are provided daily to a large state-wide group of elected and appointed leaders, public health directors, healthcare leaders and other agencies making critical planning and resource decisions. It has been useful to guide change and measure real time impact for specific communities, regions and statewide allocation of testing, PPE, ventilators and other resource allocation and workforce considerations as well as the myriad of time sensitive public health responses and policy decisions. The daily updates provide real-time early indicators of acceleration and deceleration trends that require timely attention. The PRAM analysis tool also clearly demonstrates the importance of both a regional and statewide approach to the allocation of critically important resources and the decisions regarding the application of non-pharmacological interventions (NPIs). We continue to assess the utility of the PRAM analytic tool in Nebraska and in other communities with significantly different patterns of pandemic spread and recovery.

### Data and Programming Code Availability

Nebraska Electronic Disease Surveillance System data availability is subject to the approval of the Nebraska Department of Health and Human Services: 301 Centennial Mall South, Lincoln, Nebraska 68509.

Knowledge Center data availability is subject to the approval of the Nebraska Emergency Management Agency: 2433 NW 24^th^ Street, Lincoln, Nebraska 68524-1801.

Programming code is available by contacting Chris Wichman at 984375 Nebraska Medical Center, Omaha, Nebraska 68198-4375.

## Data Availability

Nebraska Electronic Disease Surveillance System data availability is subject to the approval of the Nebraska Department of Health and Human Services: 301 Centennial Mall South, Lincoln, Nebraska 68509.
Knowledge Center data availability is subject to the approval of the Nebraska Emergency Management Agency: 2433 NW 24th Street, Lincoln, Nebraska 68524-1801.
Programming code is available by contacting Chris Wichman at 984375 Nebraska Medical Center, Omaha, Nebraska 68198-4375.

## Competing Interest Statement

The authors have declared no competing interest.

## Funding Statement

No external funding was received for this work.

## Author Declarations

The authors have nothing to declare.

## Footnotes

i The benchmark for percent positive per day is based on the average percent positive per day reported in South Korea, Germany and Iceland when the COVID outbreak was considered contained.

ii The benchmark for deaths per day is based on the assumption that no more than 2.5% of cases will end up in death.

iii The capacity limit for ventilators was set based on the tendency for ventilation to lag entry into the ICU.

